# Are men who smoke at higher risk for a more severe case of COVID-19 than women who smoke? A Systematic Review

**DOI:** 10.1101/2020.06.18.20135111

**Authors:** Aoife Rodgers, Emilie Kruke Indreberg, Lenah Alfallaj, Manasi Nadkarni, Zubair Kabir

## Abstract

**Background:** The novelty of the Covid-19 pandemic is reflected in the lack of literature available for the impact of smoking on the intensity of the COVID-19 clinical manifestations. Our study tries to address this gap.

**Method:** Six cohorts from China were analysed and a crude odds ratio was manually calculated.

**Results:** Patients with a smoking history were approximately 2 times (95% CI= 1.036-1.883) as likely to suffer from severe clinical manifestations of COVID-19 compared to patients without a smoking history. A higher percentage of males suffer more severe symptoms of COVID-19 in comparison to females, but this could be associated with the gender specific smoking trends observed in China.

**Conclusion:** The gender specific smoking trends could be associated with the increased severity of COVID-19 disease manifestations in the male population.

## BACKGROUND

Coronavirus, belonging to the Coronaviridae family, has been previously seen before and is known to cause a human illness with symptoms similar to the common cold (1). Coronavirus has been shown to be a zoonotic disease in nature, and in the case of COVID-19 originating from bats (2). In the last decade, this is the third deadliest coronavirus pandemic (2). In 2003, there was an outbreak of coronavirus known as Severe Acute Respiratory Syndrome (SARS) and in 2012 another strain of coronavirus known as Middle-East Respiratory Syndrome (MERS) emerged (3,4). A more recent strain of coronavirus has emerged named SARS-COV-2 and is generally referred to as COVID-19 (Coronavirus disease 2019). COVID-19 began in Wuhan, China in December 2019 (5). Within a month of its appearance, the World Health Organization had declared COVID-19 as a global pandemic (5). To date, there have been 3,110,696 declared cases with 215,231 deaths (6) and within the past 5 months, over 100 countries have been infected with COVID-19 (7). Due to the novelty of COVID-19, there is limited data available on the characteristics associated with COVID-19 (5).

Most human Coronaviruses cause mild symptoms. However, a subgroup of coronaviruses is causing severe outcomes. Severe Acute Respiratory Syndrome (SARS) and Middle-East Respiratory Syndrome (MERS) have a death rate of 10% and 37% respectively (8). The mortality rate for COVID-19 is almost 7% based on the current numbers (6). Shortness of breath (9), fever and cough are the main symptoms for the three coronaviruses, including COVID-19, leading to a lower respiratory infection that can be life-threatening to vulnerable groups such as to the elderly and to people with underlying medical conditions (10,11). This leads to respiratory failure and multiorgan failure (12). Cases of COVID-19 are divided into four groups mild, moderate, severe and critical pneumonia (11).

To date, there is limited data on the role smoking has to play in the progression of COVID-19, however, smoking has been identified as a risk factor for disease progression (5,13). A previous meta-analysis involving thirteen studies was carried out and determined smoking to be a risk factor in disease progression for patients with COVID-19 (13). Similarly, in the previous MERS outbreak, smokers were identified as having a higher mortality rate in comparison to non-smokers (5). In more recent literature, a systematic review of five papers in China found that smokers were 1.4 times more likely to have more severe symptoms of COVID-19 and 2.4 times more likely to be admitted to an ICU, need mechanical ventilation or died in comparison to non-smokers (RR = 2.4, 95% CI: 1.43-4.04) (5).

According to another meta-analysis that is done on 39 studies, males had more severe outcomes in comparison to females, as well as the mortality rate (11). It is not fully understood yet, however, it can be explained with the presence of estrogen is a protective factor. Also, it is linked that smoking is considered a risk factor for adverse outcome and the number of men smoking is higher than women.

Smoking prevalence can vary by country to country however in general, the smoking prevalence is higher among males than females (14),(15). It has been highlighted that developing countries and developed countries are similar in their male smoking mean (32% and 30.1% respectively) however, females in developed countries have a higher prevalence of smoking in comparison to females in developing countries (17.2% vs. 3.1%) (14). The World Health Organization’s Global Adult Tobacco Survey 2018 supports the statement males have a higher prevalence of smoking compared to females (15.6% vs 12.0%) globally (15). Due to these trends observed in the smoking difference between genders, it could lead to an indirect association between the severity of COVID-19 between genders.

## RESEARCH QUESTION

Are men who smoke at higher risk for a more severe case of COVID-19 than women who smoke?

## METHODOLOGY

### Search strategy and Selection criteria

A search was conducted in the PubMed database for studies published between January 1, 2020, to April 27, 2020. The Zotero 5.0.85 software was used to manage the selected studies. A comprehensive search was designed for the purpose of this study (Appendix 1). No language restrictions were placed on this search. Only full-text studies were included in the evidence synthesis. The studies identified by this search were further analysed for their eligibility. Eligible studies were those that demonstrated the characteristics presented in Table 2. The flow for the number of studies included in the evidence synthesis is given in Figure 1.

**Table 1:** Question in PICO format.

|  |  |
| --- | --- |
| Population | All COVID-19 cases |
| Intervention | Male smokers |
| Comparison | Female smokers |
| Outcome | Severe symptoms of COVID-19 |

**Table 2:** The inclusion and exclusion criteria used in the report.

|  |
| --- |
| **Inclusion** |
| • Cross-sectional studies, surveys, cohort studies or case studies conducted in China<br>• Studies that described the epidemiological and/or clinical characteristics of the coronavirus infection<br>• Studies that described the prevalence of smoking in the sample population, either as a whole or based on gender |
| **Exclusion** |
| • Preprints, reviews, editorials and letters<br>• Studies that provided no data about the smoking status of the sample populations affected by the coronavirus infections<br>• Studies with less than 100 participants |

**Figure 1:**
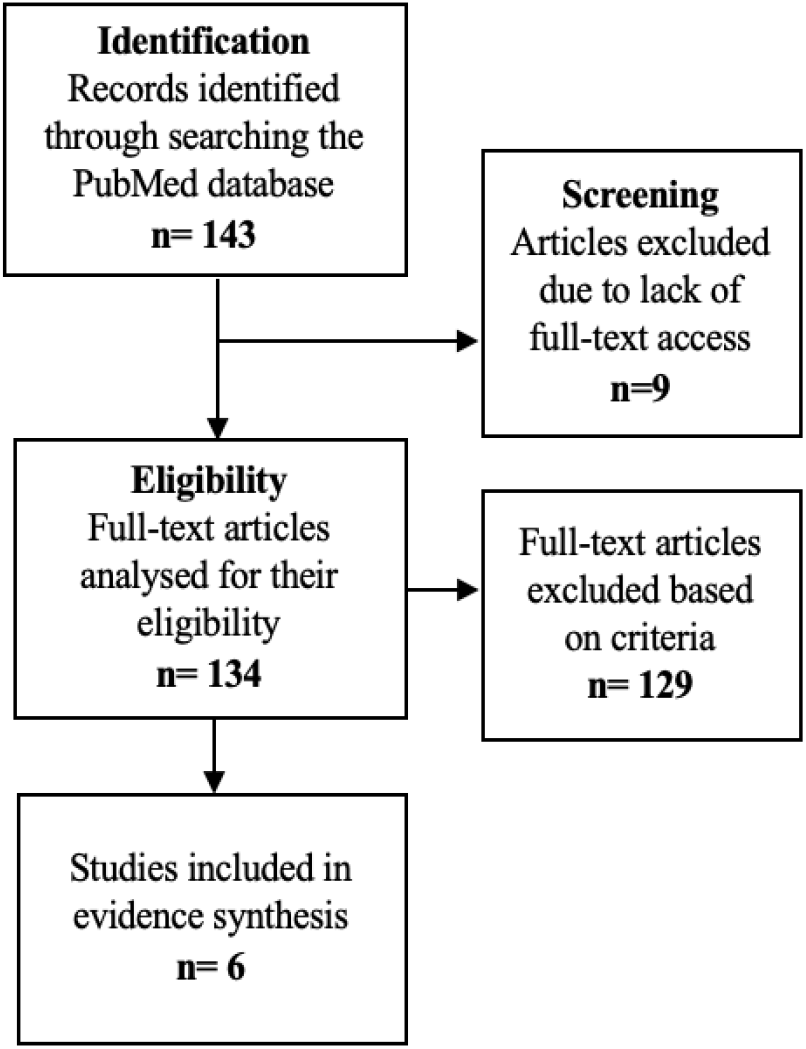
Flowchart for selecting the studies included in the evidence synthesis.

### Synthesis of evidence

The final number of included studies to synthesise for evidence was six (16,17,18,19,20,21). The six studies were mined for the following components: the total number of cases, the number of cases in males versus females, the gender stratified severity of the cases and the history of smoking (Table 3). The severe cases were those listed as severe cases, on mechanical ventilation or had died from the virus in each paper. Each study was also analysed to determine its quality. The quality of the included studies was assessed using the “Newcastle-Ottawa Quality Assessment Form for Cohort Studies”. Depending on the number of stars in each of the three domains (selection, comparability and outcome) the quality was considered good, fair or poor (22). Overall the quality for each study was considered good with low risk of bias (Appendix 4).

**Table 3:**
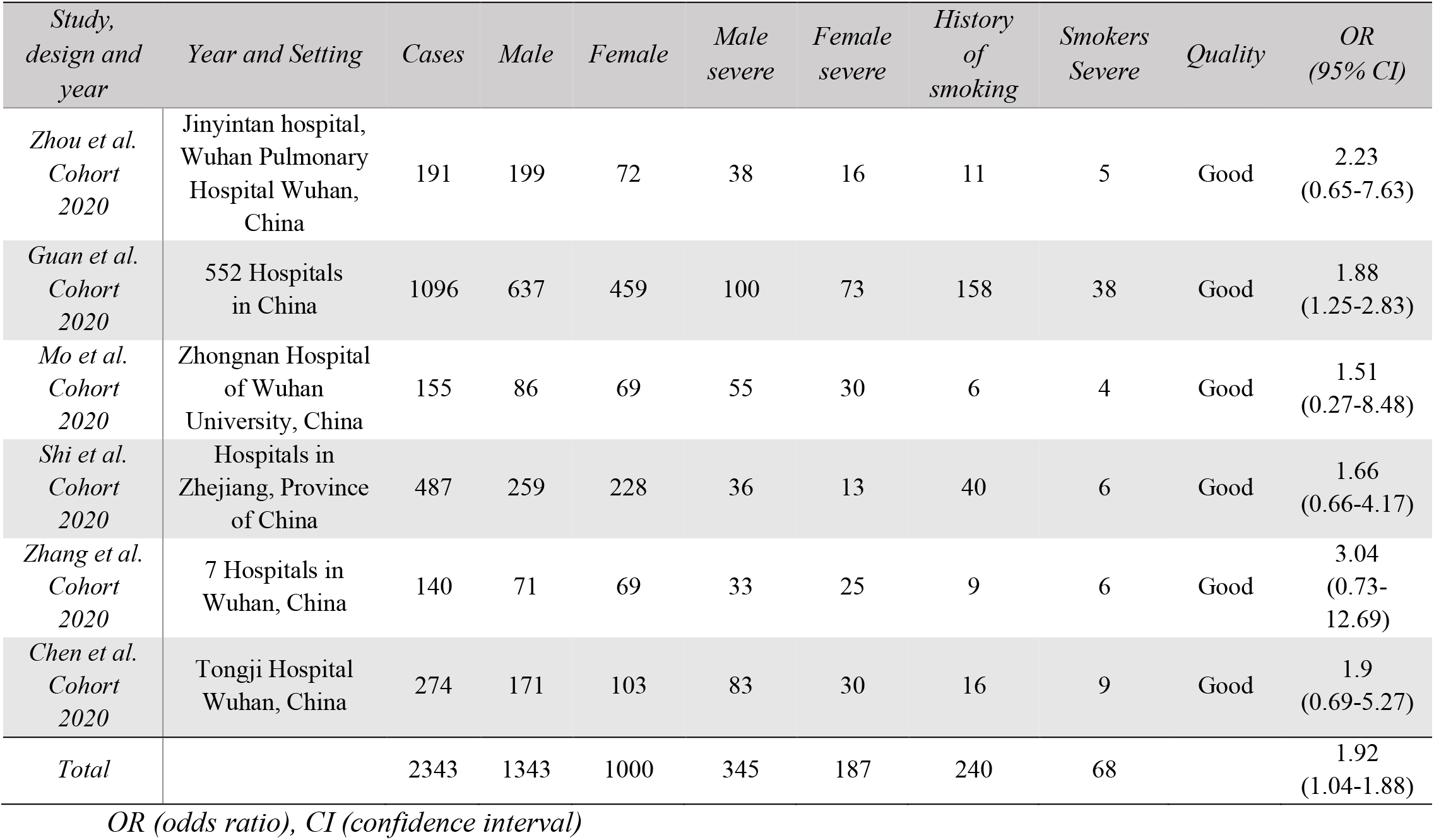
Overview of the six studies included in the review.

### Analysis

The data extracted from the studies (Table 3) was used to manually calculate a Crude Odds Ratio (COR) (Appendix 2). The mean COR was calculated as an overall estimate of the risk of severe clinical manifestations for patients with a history of smoking. The 95% Confidence Interval (95% C.I.) was estimated using the online OpenEpi tool Version 3.01.

## RESULTS

## DISCUSSION

Due to the novelty of COVID-19, there is a lack of evidence on the impact of gender-stratified smoking on disease progression. In order to elucidate the presence of this association, this paper analyzed six specific cohort studies within China to synthesize the relevant evidence. The rationale for papers only chosen within China is due to COVID-19 originating in China. China in turn, has an extensive amount of research completed on COVID-19 in comparison to any other countries worldwide. The analysis was carried out to identify whether gender differences in smoking have an effect on the severity of the symptoms on the current coronavirus strain, COVID-19, by gathering data and calculating the odds ratios for the selected cohorts. The analysis of the data extracted from these studies indicated that patients with a smoking history were approximately 2 times (OR=1.92; 95% CI= 1.036-1.883) as likely to suffer from severe clinical manifestations of COVID-19 compared to patients without a smoking history. However, there was a lack of evidence available on the differences in gender and their smoking rates. From the six cohorts in China, the smoking data for a total of 1343 males and 1000 females was analyzed. A higher proportion of males suffer from more severe symptoms of COVID-19 within China, as observed in the six cohorts where 158 more males suffered from severe cases (25%) in comparison to severe female cases (18.7%). Simultaneously, it was observed that males had a higher prevalence of smoking in comparison to females both within China (54% male smokers and 2.6% female smokers) (23). This observation supports our hypothesis that a higher proportion of men suffer from severe manifestations, as compared to females, due to the higher prevalence of smoking amongst men in China. Similarly, other countries badly affected by COVID-19, such as Italy have found that males (22.8%) are more likely to smoke in comparison to females (14.9%) (24).

Furthermore, it was observed that within the Italian population, 58% of the COVID-19 cases were male cases (25). However, in the cohort studied in Italy (Appendix 3), it was observed that patients with a smoking history were 0.35 times less likely to exhibit severe symptoms of Covid-19 than those that had no smoking history (26). Considering the fact that this is the only cohort outside China, this could indicate that further studies are needed to identify whether there are any confounders that could impact this potential association.

Previous strains of coronavirus outbreaks such as SARS outbreak in 2003 and MERS outbreak in 2012, determined a significantly higher death rate among males (21.9% vs 13.2%). The receptor used by the previous SARS-CoV viral strain to enter the host cells is the angiotensin-converting enzyme-2 (ACE2) receptor (27). It has been hypothesized that this receptor could be used as the novel adhesion molecule by the current SARS-CoV2 (COVID-19) strain. Smoking has been associated with the upregulation of this receptor molecule, thus increasing the susceptibility to more severe clinical manifestation of the disease. The use of new electronic smoking devices such as electronic cigarettes has also been hypothetically associated with this upregulation (27). This receptor upregulation and gender difference in smoking rates could help explain the higher proportion of severe cases observed in males as opposed to females, in several countries.

A similar gender-specific smoking pattern was observed in Ireland, where a study carried out by the Health Service Executive (HSE) Healthy Ireland found 24% of the male population to be smokers as opposed to 21% observed in the female population (28). Thus, indicating that Irish males are more vulnerable to severe COVID-19 cases than Irish females. As observed in Italy, a cohort study could demonstrate a negative association between smoking and severity of Covid-19 (26). This indicates that further research is necessary before any association is determined in Ireland. However, while there are no studies to support the evidence, data published by the Health Protection Surveillance Centre, states that although males account for only 46% of confirmed cases, they account for 71% of deaths, which is a severe outcome (29). This observation has an essential public health implication, as it could be used as the driving force for an amendment in the existing Irish Smoking Ban. In 2004, Ireland imposed a blanket ban on smoking in enclosed public spaces as well as workplaces. But this ban did not extend to open public spaces, such as parks, beaches, playgrounds, and campuses, or include the use of electronic cigarettes (30). In order to reduce the smoking associated gender specific disease burden, amending the current smoking ban to restrict smoking in open public places and ban the use of e-cigarettes might be necessary to reduce the incidence of severe cases of COVID-19 in the Irish population, thus decreasing the burden on the healthcare services. This amendment could also aid in achieving the 2025 targets of less than 5% smoking prevalence in Ireland (28).

## LIMITATIONS

The novelty of the Covid-19 pandemic is reflected in the lack of literature available for the impact of smoking on the intensity of the COVID-19 clinical manifestations. While our study tries to bridge this gap, it also possesses certain limitations. A meta-analysis could not be performed due to the lack of studies that matched the inclusion criteria, as less than 10 papers were selected for this review. The methods used to collect data related to smoking history varied across the studies, thus introducing heterogeneity, and, in cases of self-reported data, recall bias. While the selected papers provided gender-stratified data for the severity of cases, they did not provide the gender-stratified data for smoking. Therefore, the studies had to be supported by the gender specific smoking trends observed in the population of China to elucidate the association observed in this review. Furthermore, there was no data collected to determine whether the smoking history was recent. Thus, a concrete cause-effect relationship could not be determined on the basis of the available data.

## CONCLUSION

The analysis of the data extracted from six cohorts in China, as well as the supplementary data regarding the gender differences in smoking, indicated that smoking could be associated with the increased severity of COVID-19 disease manifestations in the male population, in comparison to the female population. Due to the similarity in smoking trends observed in other countries such as Italy and Ireland, this gender specific smoking associated distribution of severe COVID-19 cases might also be observed in these populations. Thus, an amendment in the Irish Smoking Ban, which restricts smoking in open public places such as parks, beaches, playgrounds, and campuses and a ban on the use of e-cigarettes, may help curb the incidence of severe cases of COVID-19 in the Irish population, thus decreasing the burden on the healthcare services. However, the lack of concrete gender-specific smoking data within the cohort studies suggests that further research is necessary to verify this hypothesis.

## Data Availability

Data can be available on request.

## Ethical Considerations

No personal data were used, therefore no ethical approval was sought

## Conflicts of Interest

No conflicts of interest

## Funding

No funding was available to this study. This was a part of Master of Public Health (MPH) student project.

## APPENDIX 1

Search term for evidence synthesis

| Search Number | Search Term |
| --- | --- |
| 1 | “coronavirus OR COVID-19 OR SARS OR severe acute respiratory syndrome” |
| 2 | “smoke OR smoking OR risk factor OR characteristics” |
| 3 | “smoke OR smoking OR risk factor OR characteristics” AND “sex OR gender OR men OR male OR women OR female” |
| 4 | 1 AND 2 AND 3 |

## APPENDIX 2

Crude Odds Ratio (COR) COR= (aXd)/(bXc)

The summarized data in each cell

|  |  |
| --- | --- |
| a | Number of cases that reported a smoking history as well as severe clinical manifestation |
| b | Number of cases that reported a smoking history but did not report a severe clinical manifestation |
| c | Number of cases that did not report a smoking history but reported severe clinical manifestation |
| d | Number of cases that neither reported a smoking history nor any severe clinical manifestation |

## APPENDIX 3

COVID-19 comparison

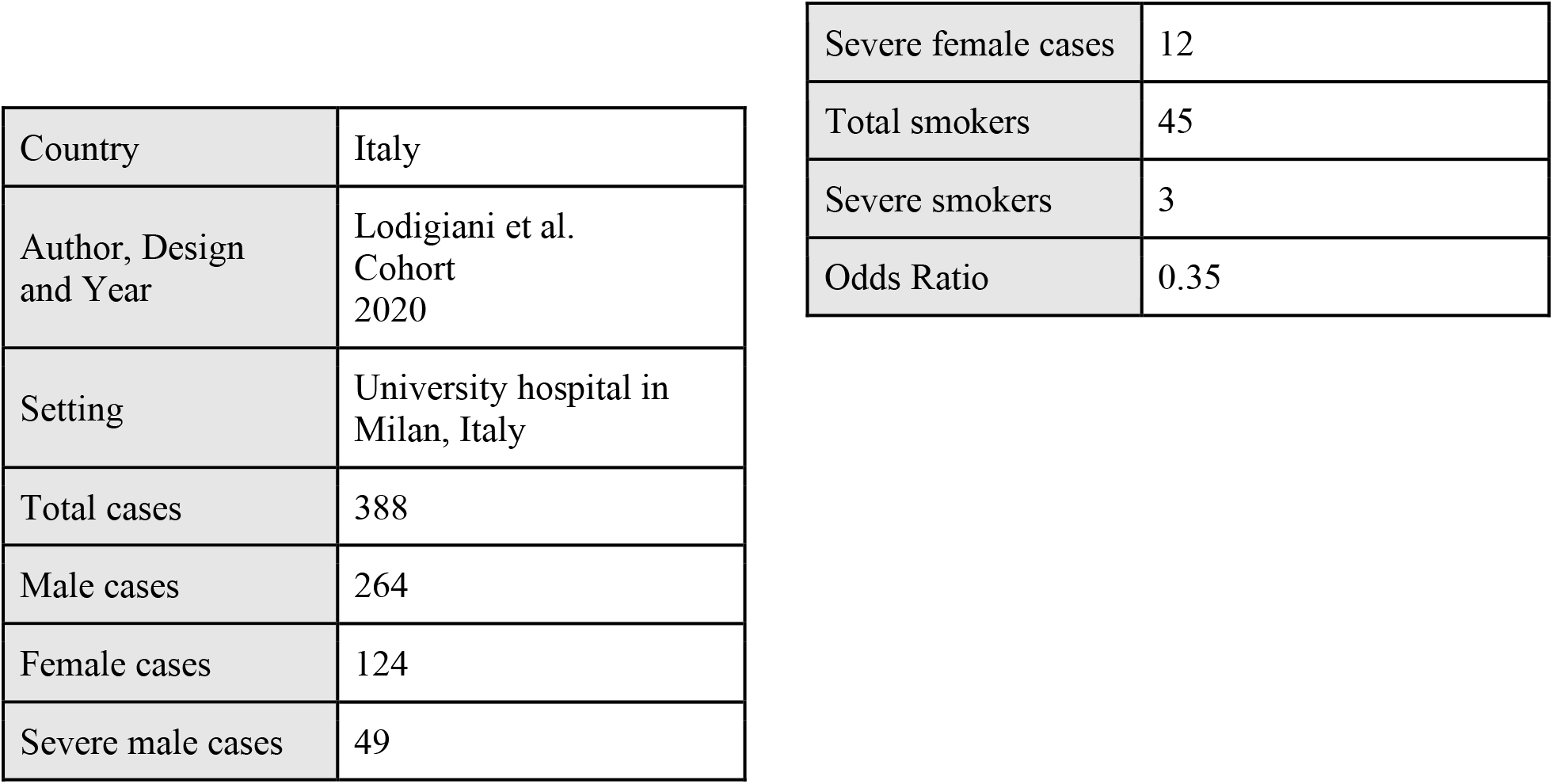

|  |  |
| --- | --- |
| Country | Italy |
| Author, Design and Year | Lodigiani et al.<br>Cohort<br>2020 |
| Setting | University hospital in Milan, Italy |
| Total cases | 388 |
| Male cases | 264 |
| Female cases | 124 |
| Severe male cases | 49 |
| Severe female cases | 12 |
| Total smokers | 45 |
| Severe smokers | 3 |
| Odds Ratio | 0.35 |

## APPENDIX 4

Newcastle-Ottawa Quality Assessment for Cohort Studies

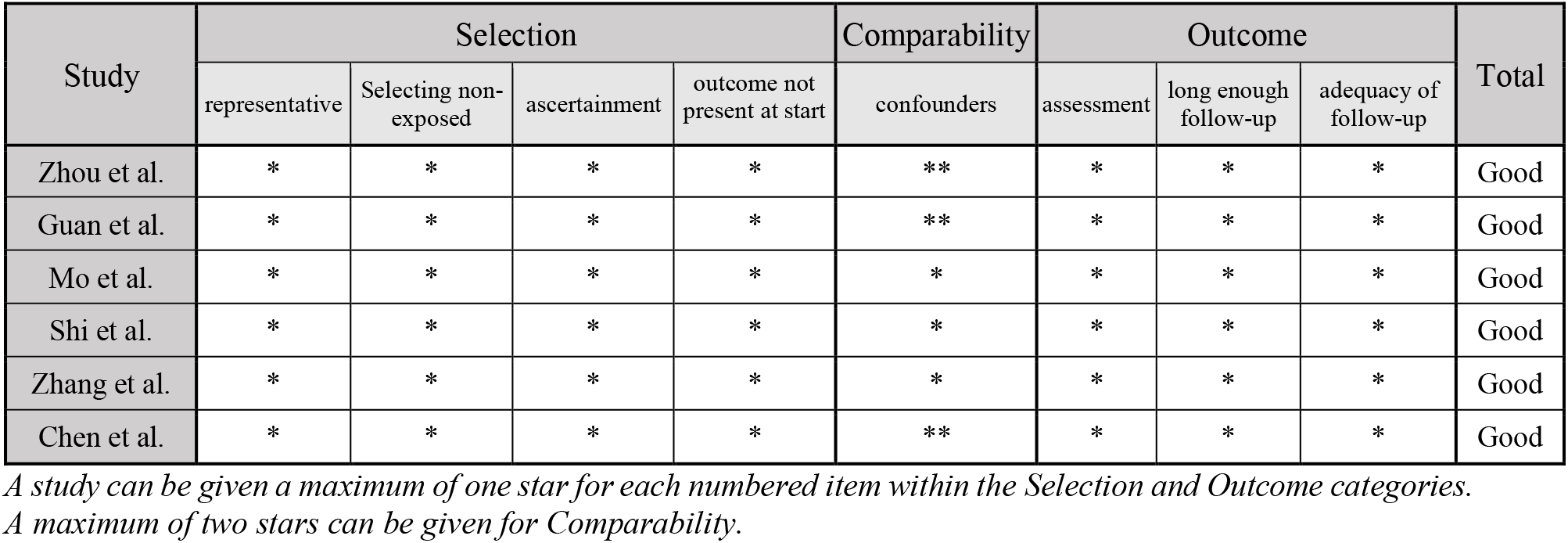

## Notes

### Competing Interest Statement

The authors have declared no competing interest.

### Author Declarations

No ethical approval was necessary

